# Antibody response to SARS-CoV-2 infection and BNT162b2 vaccine in Israel

**DOI:** 10.1101/2021.07.07.21259499

**Authors:** Guy Shapira, Ramzia Abu Hamad, Chen Weiner, Nir Rainy, Reut Sorek-Abramovich, Patricia Benveniste-Levkovitz, Adina Bar Chaim, Noam Shomron

**Affiliations:** Sackler Faculty of Medicine, Tel Aviv University, Tel Aviv, Israel; Shamir Medical Center, Be’er Yaacov, Israel

**Author notes:** Correspondence should be sent to or. Equal first authors (Guy Shapira, Ramzia Abu Hamad). Equal last authors (Adina Bar Chaim, Noam Shomron).

## Abstract

Neutralizing antibodies targeting the Spike protein of severe acute respiratory syndrome coronavirus 2 (SARS-CoV-2) block viral entry to host cells, preventing disease and further spread of the pathogen. The presence of SARS-CoV-2 antibodies in serum is a reliable indicator of infection, used epidemiologically to estimate the prevalence of infection and clinically as a measurement of an antigen-specific immune response. In this study, we analyzed serum Spike protein-specific IgG antibodies from 26,170 samples, including convalescent individuals who had coronavirus disease 2019 (COVID-19) and recipients of the BNT162b2 vaccine. We find distinct serological patterns in COVID-19 convalescent and vaccinated individuals, correlated with age and gender and the presence symptoms.

## Introduction

Severe acute respiratory syndrome coronavirus 2 (SARS-CoV-2), which is the causative pathogen of the coronavirus disease 2019 (COVID-19) pandemic, has rapidly spread worldwide, causing millions of deaths, massively impacting the economy and society^1^. The BNT162b2 vaccine, which consists of two doses of modified SARS-CoV-2 mRNA delivered in lipid nanoparticles, was proven to be effective in prevention of COVID-19 both in randomized clinical trials and nationwide, mass-vaccination settings^2,3^.

The humoral immune response to infection and vaccination works by producing acute-response antibodies in specialized cells and generating long-term memory B cells, which mediate a recall response in case of future exposure to the antigen^4^.

The presence of IgG antibodies in the serum that are specific to the receptor binding domain (RBD) of the SARS-CoV-2 Spike protein is a reliable and widely used marker of past infection^5^. Serological testing is used to assess the prevalence of infection within a population^6^, as well as being a useful marker for disease severity and progression^7–9^.

In this study, we measured the concentration of SARS-CoV-2 RBD-specific serum antibodies in an Israeli cohort, containing individuals vaccinated with the BNT162b2 vaccine, patients recovering from COVID-19 and others who tested negative to SARS-CoV-2-specific serum antibodies (seronegative). We provide a comprehensive comparison between the serological antibody profile arising from vaccines or infection-acquired antibodies in symptomatic and asymptomatic individuals, and explore their relationship with clinical and demographic parameters.

## Results

### Study population

A total of 26,170 samples were collected between November 8, 2020 and March 5, 2021, 3,513 (13.4%) were recipients of the BNT162b2 vaccine (this includes first and second dose recipients unless noted otherwise); 1,653 (6.3%) were recovering from symptomatic, PCR-verified COVID-19; and, an additional 21,085 (80.5%) were sampled in national routine checkups and did not experience any COVID-19 symptoms (Table 1).

**Table 1:** Cohort demographic characteristics, by groups.

| Characteristics | Population before vaccination campaign (N=8,078) |  | Population after vaccination campaign (N=12,924) |  | Recovering from symptomatic COVID-19 (N=1,652) |  | Vaccinated (N=3516) |  |
| --- | --- | --- | --- | --- | --- | --- | --- | --- |
|  | Range | Mean±SE | Range | Mean±SE | Range | Mean±SE | Range | Mean±SE |
| Age | 1-100 | 40.48±0.19 | 4-93 | 37.86±0.12 | 6-88 | 40.67±0.39 | 15-99 | 52.9±0.27 |
| S1/S2 Antibodies (AU/mL) | 4.9-374 | 20.94±0.37 | 4.9-800 | 78.56±1.74 | 4.9-800 | 65.32±2.6 | 4.9-800 | 231.96±3.68 |
| Male gender – no. (%)* | 4862 (61%) |  | N/A |  | 323 (42%) |  | 1041 (44%) |  |

\*Observations with missing gender excluded.

Seropositive samples taken before the beginning of the 20/12/2020 Israeli national vaccination campaign (N=8,078) are from unvaccinated individuals suspected to have had asymptomatic COVID-19. IgG antibodies against S1/S2 SARS-CoV-2 antigens were measured from serum samples, yielding AU/mL values in the 4.9-800 range, with 15 being the cutoff for a positive result (see methods).

### Comparison of SARS-CoV-2 and BNT162b2-elicited antibody response

Vaccinated individuals had the highest antibody levels, nearly three times higher than that of convalescent individuals recovering from symptomatic COVID-19 (Wilcox P < 0.001). Similarly, the vaccination campaign caused a population-wide increase in antibody levels, on average almost four times higher than samples taken before the campaign (Wilcox P < 0.01) (Fig 1) (Table 1).

**Figure 1:**
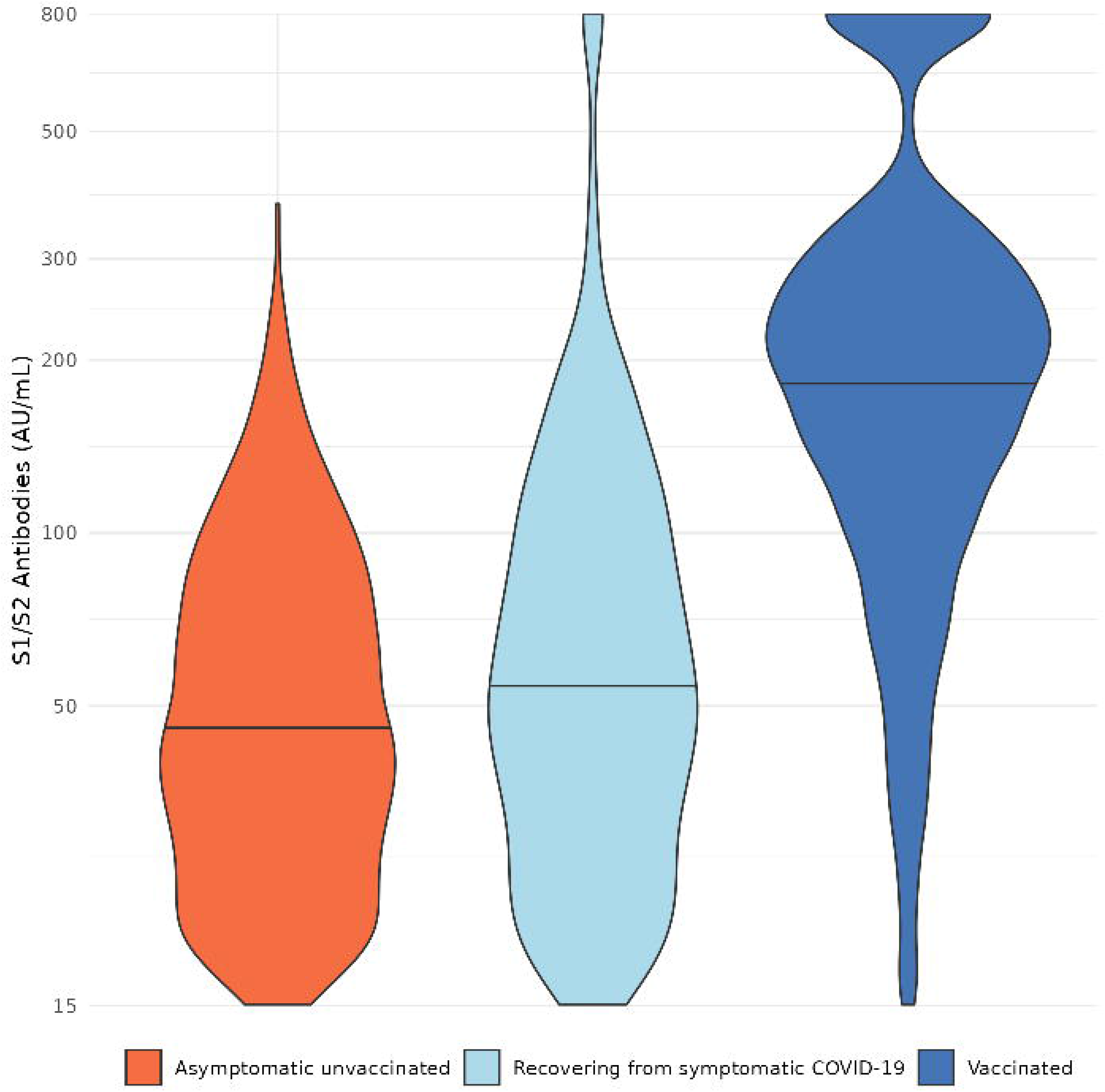
Violin plot of serum S1/S2 antibodies levels (AU/mL) of seropositive samples from symptomatic and asymptomatic unvaccinated individuals and vaccine recipients. The horizontal lines mark median values per group.

Nearly all vaccinated individuals were seropositive six days or more after the second dose (99.4%), while patients recovering from symptomatic COVID-19 were only mostly seropositive (75.7%) (RR=1.31; Chisq P<2.9e-36) (Fig 2).

**Figure 2:**
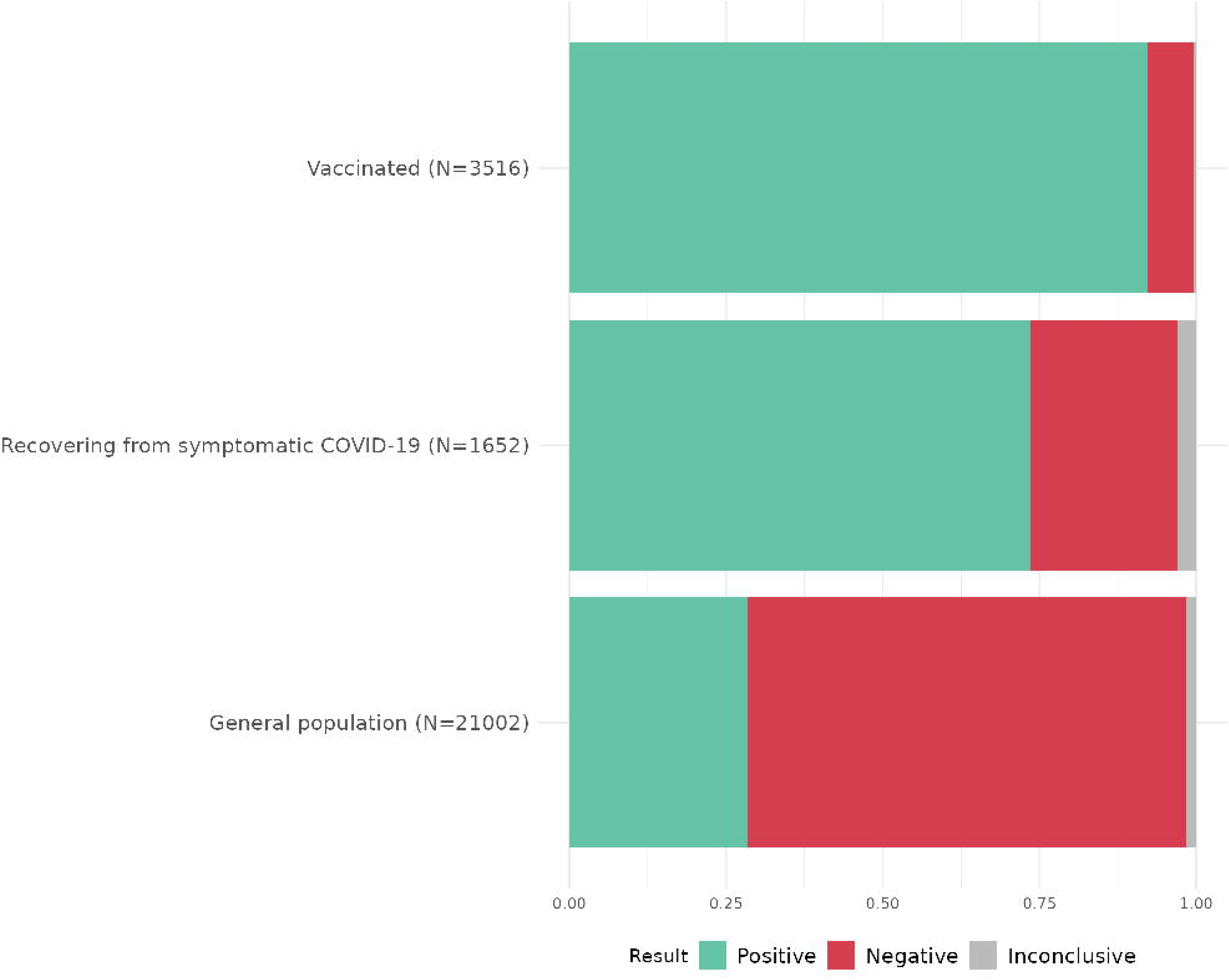
Bar plot of serum assays by prevalence of seropositivity (fraction) in a general asymptomatic population sample, unvaccinated individuals recovering from symptomatic COVID-19 and vaccine recipients. Assay results too close to the seropositivity cutoff were marked inconclusive.

We observed an inverse correlation between age and antibody concentration in vaccinated individuals (Pearson=-0.1 P=2.9e-9). Seropositive patients recovering from symptomatic COVID-19 were slightly, albeit significantly older than their seronegative counterparts (41.5±0.4 and 38.3±0.7, respectively; Wilcox P=0.0004) (Fig 3).

**Figure 3:**
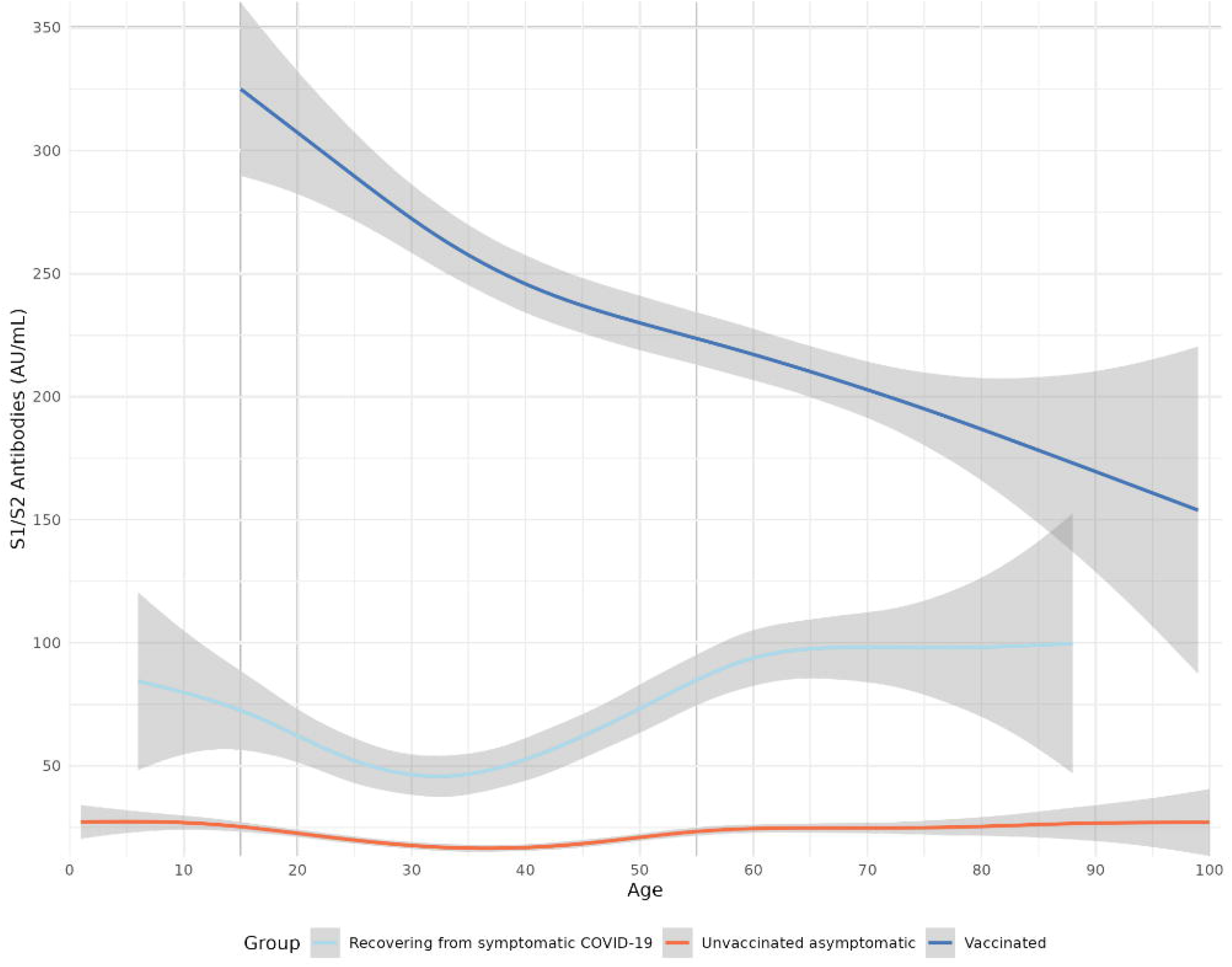
Levels of serum S1/S2 antibodies (AU/mL) by age, in symptomatic and asymptomatic unvaccinated individuals and vaccine recipients.

### Dynamics of BNT162b2-elicited antibody response

Antibody levels of vaccinated individuals are dynamic, displaying a distinct, two-phase pattern of rapid increase in the seroconversion phase, followed by stabilization or decline in the latter phase.

In the first 10 days following the second vaccination, IgG antibody levels rapidly increase across all age groups, with the young-adult group having a significantly larger and steeper increase (Wilcox P=0.0003; when comparing ages [18,44] to (55,83]) (Fig 5). IgG antibodies reach a stable plateau in days 10-12, then, the trend shifts to being mostly stable for adults in the 18-44 age groups (Wilcox P=0.2), and significantly declining in the older 50-86 age group (Wilcox P=0.0004).

**Figure 4:**
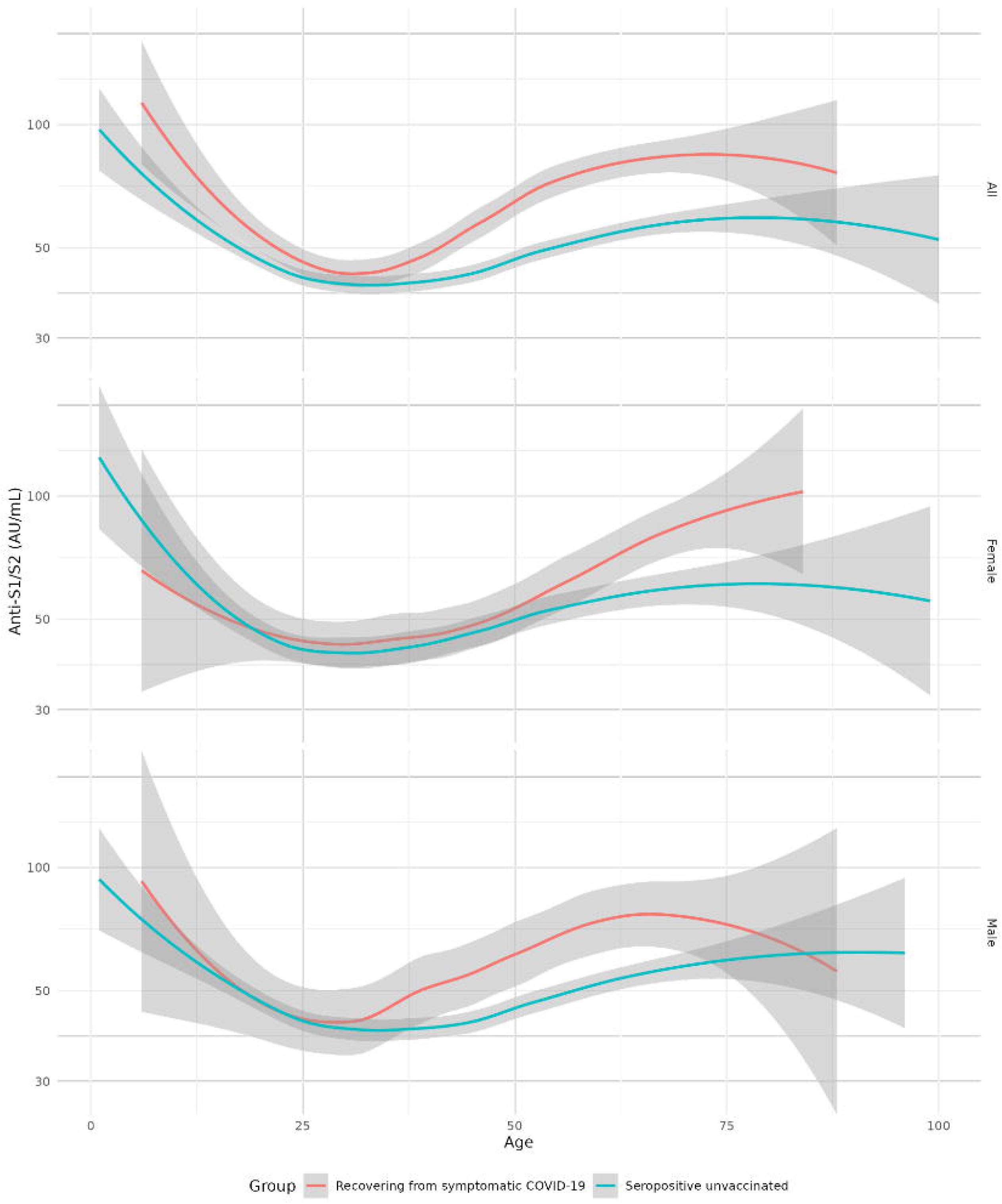
Levels of serum S1/S2 antibodies (AU/mL) in unvaccinated convalescent individuals, split to individuals recovering from symptomatic COVID-19 and a seropositive unvaccinated group suspected to have had asymptomatic COVID-19.

**Figure 5:**
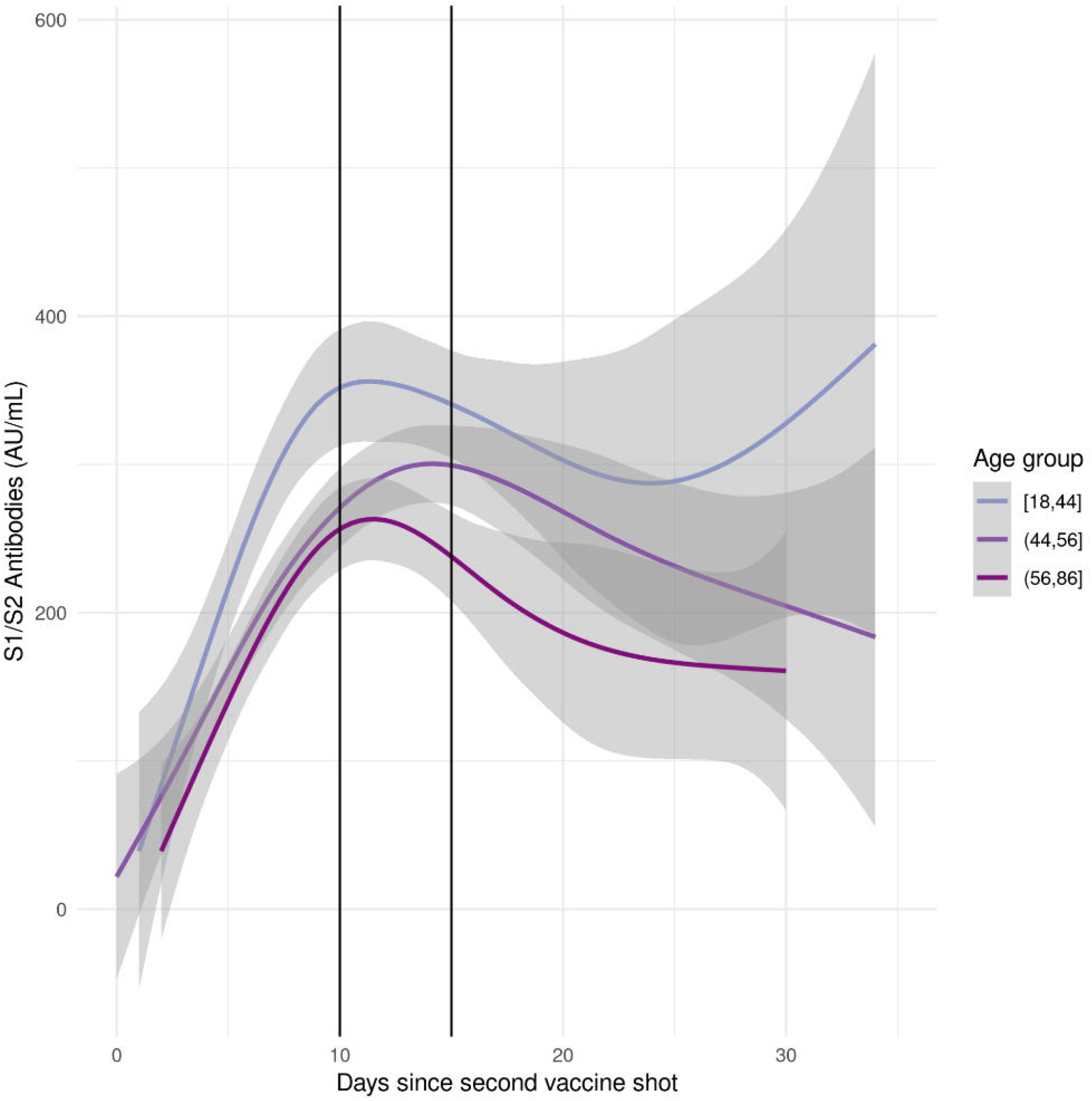
S1/S2 Serum antibodies in the days following the second vaccine shot, stratified by age groups. Vertical black lines mark boundaries between the stages of antibody response: phase 1 (increase), plateau and phase 2 (stabilization/decline).

It is important to note that despite the sharp decline (261±18.2 in plateau, declining to 180±7.7 in the second phase), patients in the 56-86 age range maintained antibody levels that are well above the seropositivity detection threshold (15 AU/ml).

### COVID-19 and convalescent antibody concentration

We compared results from seropositive convalescent individuals recovering from symptomatic COVID-19, with seropositive unvaccinated individuals suspected to have had asymptomatic COVID-19 at some point.

There is a distinct U-curve association between age and antibody concentration in both groups, starting with high concentrations in infants, then declining to a stable low level in young adults, then rising again around age 35 (Fig 3).

Regardless of age, individuals recovering from symptomatic COVID-19 had significantly higher antibody levels compared to asymptomatic ones (Kruskal P=2.3e-09).

Regarding gender, with males the difference between recovered and the seropositive unvaccinated group is highly significant (Kruskal P=0.0005), while in females, it is weak and only marginally significant when age is unaccounted for (Kruskal P=0.06). The difference in antibody levels in males becomes significant after age 30 (Kruskal P=8e-6), while in females the difference is apparent much later, most notably among women aged 51 or older (Kruskal P=6e-4).

A closer examination reveals that the difference in antibody levels between the symptomatic and asymptomatic groups is generally insignificant in ages 30 or younger (Kruskal P=0.44), but becomes highly significant in ages 30 and older (Kruskal P=7.1e-11).

## Discussion

In this study, we characterized the antibody response to the BNT162b2 vaccine and SARS-CoV-2 infection, in relation to demographic and clinical parameters.

The serum anti-RBD IgG antibodies that were assayed are highly accurate markers of infection^10^ and strongly correlate with neutralizing activity^11^ and disease severity^7^, but can not be used as sole predictors of anti-SARS-CoV-2 neutralizing ability^12^. Convalescent individuals recovering from symptomatic COVID-19 typically have low plasma titres of RBD-specific antibodies, however, the antigen-specific memory B cells that facilitate the antibody response, maintain and enhance their potency for years^11,13,14^.

In agreement with previous studies, levels of IgG serum antibodies elicited by the mRNA vaccine were significantly higher than those of convalescent individuals^11,15,16^ and inversely correlated with age^17^ (Fig 3). In the month following the second BNT162b2 dose, older individuals had lower antibody levels in the seroconversion phase, followed by an earlier and steeper decline compared to younger age-groups (Fig 5). Despite decreasing quantities, the potency of the antibodies is mostly unaffected by aging^18^ and neutralizing activity is possible at much lower concentrations^11^. Taken together with the results of large-scale epidemiological studies^19,20^, the evidence currently suggests that BNT162b2 maintains its efficacy in the oldest age groups.

Among unvaccinated convalescent individuals, antibody levels were highest in young children, decreasing to stable low levels in young adulthood, then increasing again in later adulthood (Fig 4). The prevalent association of elevated antibody levels to increased COVID-19 severity is contradicted by the highest antibody concentrations belonging to the youngest and least vulnerable age-group^21,22^. Like many other aspects of the pediatric immune response to SARS-CoV-2, this phenomenon remains largely unexplained^22^.

Overall, seropositive unvaccinated individuals recovering from symptomatic COVID-19 had higher antibody levels than their asymptomatic counterparts, however, this difference was highly gender and age-dependent (Fig 4). Among children and young adults, there was a very slight, insignificant increase in antibody levels in severe cases, which was previously associated with asymptomatic and mild COVID-19^23^. In later adulthood, the antibody levels of the symptomatic COVID-19 group increased significantly more than the asymptomatic group and the gap between the two grew larger with age. Among females, the difference in antibody levels between the symptomatic and asymptomatic COVID-19 groups was the most significant in age 51 and older, while in males this separation occurs much earlier, at about 35 years of age.

Women are at significantly lower risk of developing severe COVID-19, this is mostly attributed to the immunomodulatory effect of estrogen, which serves as a protective factor^24,25^. According to our data, the disproportionate increase in antibody levels of women recovering from symptomatic COVID-19 starts at age 51, coinciding with the mean age of menopause^26^. It is therefore implied that rising antibody levels in women over the age of 51 might be the result of the menopausal drop in estrogen, which nullifies it’s immunomodulatory effect, increasing the incidence of immune dysregulation associated with the more severe and symptomatic COVID-19^27^.

Following this conclusion, it can be postulated that the early separation between the symptomatic and asymptomatic COVID-19 groups in men reflects increased susceptibility and coincides with a major COVID-19 risk factor. It is currently estimated that male testosterone levels decrease significantly by age 40^28^, roughly coinciding with the formation of a gap between symptomatic and asymptomatic men. Unlike estrogen, however, the effect of testosterone on COVID-19 pathology is less consistent and the association is not as robust^29,30^.

In conclusion, the humoral response to SARS-CoV-2 infection and vaccination is distinct and varied across age and gender. Disparities in antibody levels between the various groups are reflective of numerous under-explored phenomena with potential clinical implications, such as the discordance between symptomatic and asymptomatic COVID19 in relation to age and sex.

## Materials and Methods

### Sample collection and assays

Whole blood samples were collected into SST gel tubes using a standard technique at the patient’s house or in the hospital. Blood samples were kept at 2-8 C° degrees and transferred to Shamir Medical Center laboratory within two hours. Samples were Centrifuge for at least 10 minutes at 3000 RPM.

COVID-19 serological tests were performed using the following commercially available, FDA approved, automated immunoassays:

- The LIAISON® SARS-CoV-2 S1/S2 IgG (311450, DiaSorin, Saluggia, Italy): A chemiluminescent immunoassay (CLIA) for quantitative determination of anti-S1 and anti-S2 specific IgG antibodies using magnetic beads coated with S1 and S2 antigens. The analyzer automatically calculates SARS-CoV-2 S1/S2 IgG antibody concentrations expressed as arbitrary units (AU/ml), with a positive cutoff level of 15.0 AU/ml.
- The LIAISON® SARS-CoV-2 TrimericS IgG assay (Emergency Use Authorization, EUA) is able to identify patients diagnosed for COVID-19 by virus variants (Lineage B.1.1.7 and Lineage P.1). The analyzer automatically calculates SARS-CoV-2 S1/S2 IgG antibody concentrations expressed as arbitrary units (AU/ml), with a positive cutoff level of 15.0 AU/ml. The principal components of the test are paramagnetic particles (solid phase) coated with recombinant trimeric SARS-CoV-2 spike protein and a conjugate reagent containing an anti-human IgG mouse monoclonal antibody linked to an isoluminol derivative. The assay is intended to assess the presence of antibodies to SARS-CoV-2, including neutralizing antibodies, in human serum or plasma.

### Data processing and filtering

A global minimum and maximum values were set for the assay results, generalizing the detection range to (4.9, 800) so that it applies to both kits. All analyses were performed using R 4.1.0, on a 64 bit Linux system.

Multivariate analyses were performed using one-way ANOVA and post-hoc group-comparisons were done using the Wilcoxon test.

Figures 2-5 are visualizations of generalized linear models with 95% confidence intervals, with figure 3 being a generalized additive model (GAM) and figures 4-6 were modelled by local estimated scatterplot smoothing (LOESS).

**Figure 6:**
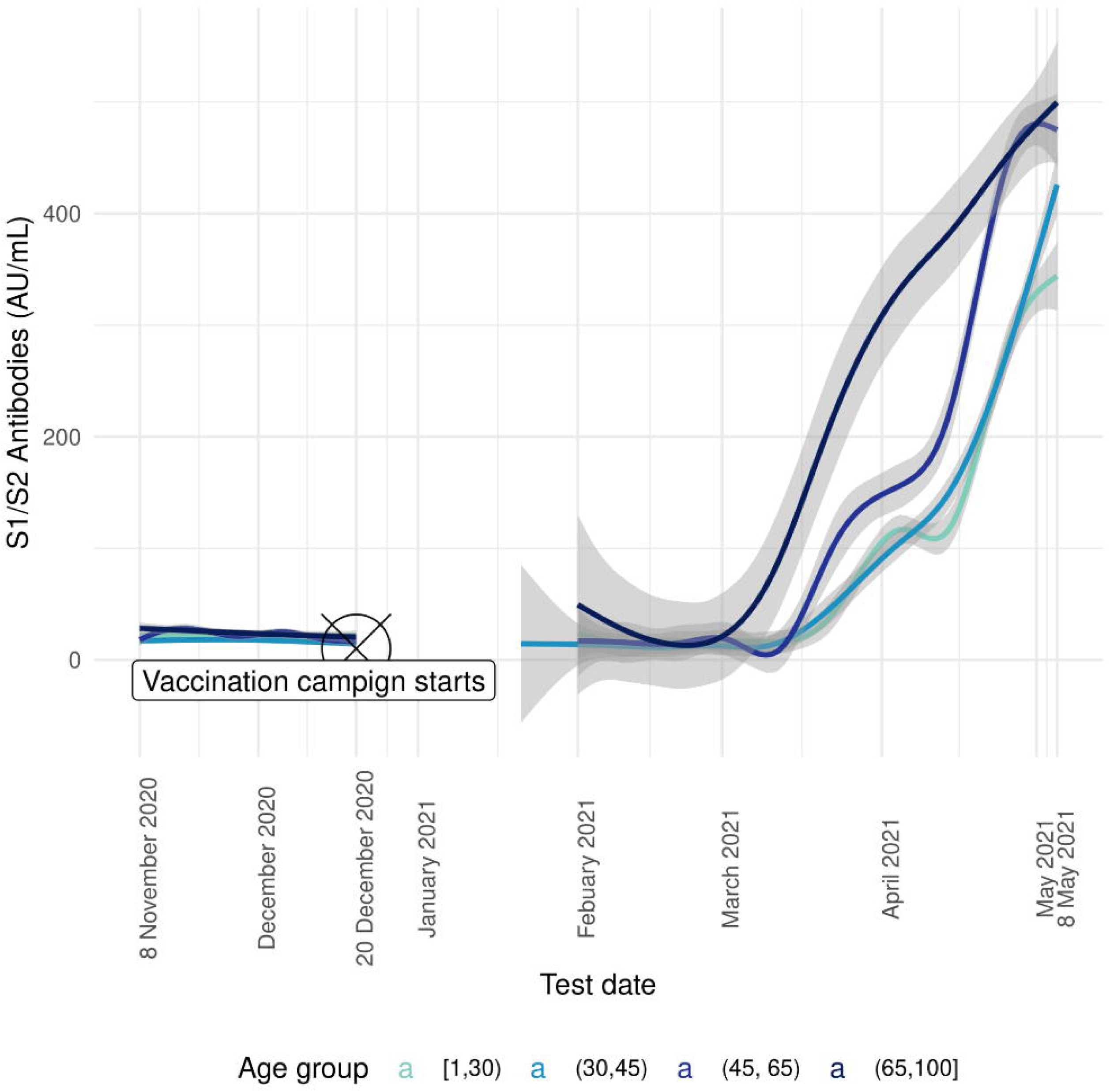
S1/S2 Serum antibody levels in the general population, stratified by age groups. The beginning of the vaccination campaign is marked, followed by a lockdown period, when no testing was performed.

## Data Availability

The data collected for this study is protected under patient confidentiality laws.

## Notes

### Competing Interest Statement

The authors have declared no competing interest.

### Funding Statement

This study was not supported by any specific grant and required no additional funding.

### Author Declarations

The study was approved by the Institutional Review Board of Shamir (Assaf Harofe) Medical Center and followed the tenets of the Declaration of Helsinki (146-21-asf).

